# Clinical Surveillance Identifies SARS-CoV-2 Outbreaks and Emergence of Novel Variants in Real-Time

**DOI:** 10.1101/2025.09.09.25335437

**Authors:** Steven C. Holland, ABCTL Diagnostic Testing and Sequencing Teams, Ian Shoemaker, Theresa Rosov, Caroline Compton, Joshua LaBaer, Efrem S. Lim, Vel Murugan

## Abstract

Monitoring community health and tracking SARS-CoV-2 evolution were critical priorities throughout the COVID-19 pandemic. However, widespread shortages of personal protective equipment, the necessity for social distancing, and the redeployment of healthcare personnel to clinical duties presented significant barriers to traditional sample collection. In this study, we evaluated the feasibility of using self-collected saliva specimens for the qualitative detection of SARS-CoV-2 infection. Following confirmation of reliable viral detection in saliva, we established a large-scale surveillance program in Arizona, USA, to enable clinical diagnosis and genomic sequencing from self-collected samples. Between April 2020 and December 2023, we tested approximately 1.4 million saliva samples using RT-PCR, identifying 94,330 SARS-CoV-2 infections. Whole genome sequencing was performed on 69,595 samples, yielding 54,040 high-quality consensus genomes. This surveillance approach enabled real-time monitoring of infection trends, outbreak detection within specific populations, and the identification of novel viral lineages over the course of the pandemic. The co-location of clinical testing and sequencing capabilities within the same facility significantly reduced turnaround time from the identification of positive cases to the generation of sequencing data. Our findings support the use of self-collected saliva as a scalable, cost-effective, and practical strategy for infectious disease surveillance in future pandemics.

## 1. Introduction

The novel coronavirus SARS-CoV-2, the causative agent of COVID19 disease, was first observed in Wuhan, China in November 2019 [1]. On January 30, 2020, the World Health Organization (WHO) declared the SARS-CoV-2 virus and COVID19 a public health emergency of international concern (PHEIC) [2]. On May 5, 2023, WHO ended the PHEIC designation of SARS-CoV-2 and COVID19 [3]. Over the three-year public health emergency, over 6*7*0 million cases of COVID19 and over 6.8 million deaths are estimated to have occurred due to SARS-CoV-2 [4].

To diagnose many respiratory infections, the recommended patient sample type is nasopharyngeal swab (NPS) or nasopharyngeal aspirate (NPA). These sample collection methods are often uncomfortable for patients and usually require collection by a trained healthcare worker, increasing the risk of virus transmission [5]. Because of these considerations and lower cost, saliva has been increasingly studied for its value in diagnosing respiratory infection [6]. Throughout the pandemic, saliva was analyzed for its ability to make qualitative, diagnostic assessment of SARS-CoV-2 infection and was found to be a reliable sample matrix [7,8].

Early in the pandemic, the US Centers for Disease Control described primer and probe sequences for the detection of SARS-CoV-2 via nucleic acid amplification tests [9]. Shortly after, independently designed commercial nucleic acid amplification tests became available for diagnostic and research use. On March 13, 2020, the TaqPath COVID-19 Combo Kit received Emergency Use Authorization (EUA) for detection of SARS-CoV-2 in nasopharyngeal swabs, bronchoalveolar lavage, mid-turbinate swabs, nasal swabs, nasopharyngeal aspirate, and oropharyngeal swabs [10]. The TaqPath COVID-19 Combo Kit contained primers and probes targeting the SARS-CoV-2 orf-1ab, S, and N genes but the oligo sequences were not publicly available. Evolution of SARS-CoV-2 increasingly led to the discovery of lineages with mutations affecting test sensitivity at one gene locus, termed gene target failure (GTF) [11,12]. On July 30, 2021, the TaqPath COVID-19 Fast PCR Combo Kit 2.0 was designed and granted EUA for use on saliva samples [13]. While exact assay designs are not publicly available, the manufacturer describes that each gene (Orf1a, Orf1b, and N) is interrogated with multiple probes sharing the same fluorescent moiety [14]. With this scheme, multiple mutations within a gene are required for target failure and the highly mutating S gene is no longer a gene target.

The SARS-CoV-2 pandemic occurred during, and further bolstered, the increased availability of next generation sequencing (NGS) capabilities. The whole genome sequencing of pathogens can inform public health guidance with regard to the detection novel and recombinant virus lineages containing mutations of health and therapeutic interest. One sequencing strategy used to increase target nucleic acid concentration in samples is tiled amplicon sequencing [15]. This strategy uses primer oligos to PCR amplify the whole target genome across two or more library pools. Like RT-PCR, tiled amplicon sequencing also relies on primer homology to create sequencing amplicons and is also susceptible to viral evolution causing the unsuccessful amplification of some amplicons. The popular ARTIC network protocol uses over 90 amplicons to amplify the approximately 30kb genome of SARS-CoV-2 and the primer scheme was updated multiple times throughout the pandemic to account for mutations in primer regions causing reduced amplification sensitivity [16].

With the increased available knowledge of whole genome evolution of the SARS-Cov-2 virus provided by NGS, the creation of naming conventions to describe novel viral phylogenetic lineages below the species level was needed. The PANGO nomenclature system was proposed and adopted to describe virus lineages which contained specific constellations of mutations and comprised their own distinct phylogenetic branches [17]. Because this nomenclature captured mostly genotypic information, in 2021, the WHO announced a naming scheme that would supplement PANGO classification in order to highlight PANGO lineages with notable phenotypic traits [18]. To facilitate public health communication, notable lineages would be designated as variants of interest (VOIs) and variants of concern (VOCs) and be given Greek letter labels depending on their transmissibility, virulence, and clinical presentation characteristics. WHO later added a variant under monitoring (VUM) designation, which aimed to raise awareness to lineages based on phylogenetic and genotypic data, before robust phenotypic data was available [19].

To assess the epidemiological state and viral evolution of SARS-CoV-2 in Arizona, USA, we established a multi-yearlong study using self-collected saliva samples. Over 1.4 million RT-PCR assays were performed on saliva samples from over 360,000 participants. Over 94,000 samples tested positive for SARS-CoV-2 infection and over 69,000 samples were subjected to NGS to obtain whole SARS-CoV-2 consensus genomes. Over the survey period, our efforts resulted in the deposition of over 54,000 SARS-CoV-2 consensus genomes into the GISAID database. Due to the robust results and the ease of sample collection, we believe self-administered saliva collection could provide an effective means for analyzing future respiratory disease outbreaks.

## 2. Materials and Methods

### 2.1 Nucleic acid extractions

Nucleic acid extractions were performed using the MagMAX Total RNA Isolation Kit (Invitrogen, Waltham, Massachusetts, USA) using a KingFisher Flex (Thermo Scientific, Waltham, Massachusetts, USA). Briefly, 200 µL of saliva sample or NP viral transfer medium (VTM) was added to 10 µL of total nucleic acid magnetic beads and 265 µL Binding Solution. For NPS, 400 µL of NPS storage solution was added to 20 µL of total nucleic acid magnetic beads and 530 µL Binding Solution. Sample binding mixes, required buffers, and wash solutions were prepared in 96-well format using a Biomek i*7* automated liquid handler (Beckman Coulter Life Sciences, Indiana, USA). With the MagMAX instrument loaded with wash solutions, diluted TURBO DNase, and elution buffers as directed by the manufacturer’s protocol, the ‘MagMAX Total’ program was run on the KingFisher Flex. Aliquots of nucleic acid extracts were used for immediate testing and the remainder stored at −80°C.

### 2.2. RT-PCR assays

Nucleic acid extracts run on the TaqPath COVID19 Combo Kit (Applied Biosystems; Waltham, Massachusetts, USA) were assayed following the manufacturer’s instructions for 384-well format. Assay plates were prepared using a custom automation method on a Biomek i*7* automated liquid handler. For saliva samples, 11 µL of nucleic acid extract was used as sample input and combined with 11 µL reconstituted Reaction Mix. For NPS, 5 µL of nucleic acid extract was used as sample input and combined with 20 µL reconstituted Reaction Mix. The RT-PCR cycling reactions were performed and analyzed on a QuantStudio7 Flex (Applied Biosystems; Waltham, Massachusetts, USA) instrument with the following cycle conditions: 1 cycle of 25°C for 2 minutes, 53°C for 10 minutes, 1 cycle of 95°C for 2 minutes, and 40 cycles of 95°C/60°C for 3 seconds/30 seconds. Assay interpretations were performed as per manufacturer’s recommendations.

Saliva samples run on the TaqPath COVID-19 Fast PCR Combo Kit 2.0 (Applied Biosystems; Waltham, Massachusetts, USA) were assayed following the manufacturer’s instructions for 384-well format. Assay plates were prepared using a custom automation method on a Biomek i*7* automated liquid handler. Briefly, 22 µL of saliva was added to 22 µL SalivaReady and incubated for 5 minutes at 62°C and then 5 minutes at 92°C. Then 14 µL of the SalivaReady reaction was combined with 6 µL of reconstituted Assay Reaction Mix. The RT-PCR cycling reactions were performed and analyzed on a QuantStudio*7* Flex (Applied Biosystems; Waltham, Massachusetts, USA) instrument with the following cycle conditions: 53°C for 5 minutes, 1 cycle of 85°C for 5 minutes, 1 cycle of 95°C for 2 minutes, and 40 cycles of 95°C/62°C for 1 second/30 seconds. Assay interpretations were performed as per manufacturer’s recommendations.

### 2.3 Limit of detection and heat stability testing of SARS-CoV-2 in saliva

Heat-inactivated SARS-CoV-2 virus particles were obtained from ATCC (Catalog #VR-1986HK). Three aliquots of virus particles were diluted to 100,000 copies/µL in a pool of SARS-CoV-2 negative saliva from 10 individuals and diluted to 3.1 copies/µL using 2-fold serial dilutions. Sample nucleic acids were extracted using the MagMAX assay and assayed for detection of SARS-CoV-2 using the TaqPath COVID19 Combo Kit. For confirmation of limit of detection, 24 samples at 100,000 SARS-CoV-2 virus particles/µL were generated and serially diluted to 12.5/µL. Sample nucleic acids were extracted using the MagMAX assay and tested for SARS-CoV-2 detection using the TaqPath COVID19 Combo Kit at concentrations 1x, 2x, 4x, 8x, 16x, and 24x the limit of detection.

### 2.5 Paired NPS and saliva sample collection

Paired saliva and NPS were collected from 148 individuals. NPS were collected by trained nurses and stored in 500 µL VTM. Saliva samples were collected by each participant. NPS and saliva samples were collected within 30 minutes of each other. NPS and saliva samples had their nucleic acids extracted and assayed for the presence of SARS-CoV-2 using the TaqPath COVID19 Combo Kit as described above.

### 2.5 Saliva sample kit creation, distribution, collection, and testing

Saliva collection kits were composed of a set of sample collection and registration instructions, an absorbent towel, alcohol wipe, a sample return bag, and a 6 ml collection tube (Micronic, Lelystad, Netherlands) labeled with a 1 ml minimum sample volume and unique tube identifier (ID). Each tube contained two identifiers consisting of a 10-digit numeric and a 5-digit alphanumeric. 5-digit alphanumeric identifier was mathematically derived from the 10-digit. Participants were requested to enter both IDs to verify the integrity of sample IDs, to make sure that the delivery of sample results to the right participant. Collection kits were placed near sample return locations and distributed to partner institutions.

Participants were recruited from partnered private organization employees, governmental organization employees, university students, and eventually their dependents and families. Recruitment was comprised of self-initiated testing and institution selected testing. Testing frequency and selection method were determined by each institution independently. Depending on availability, the participants were able to choose a supervised collection site or to drop off a sample at an unsupervised site.

From April 4, 2020, until December 2021, supervised self-collected saliva drop-off sites were available. At these locations, trained staff and nurses were available to aid in kit distribution, sample collection, and sample registration. At these locations, deposited samples were maintained at 4°C and were collected every day.

For testing after February 2021, unsupervised drop-off sites were created where participants deposited their self-collected samples in sample deposit boxes. Sample deposit boxes varied but generally were a lockable container having a lid with a hole for sample return. Over the course of the survey period, approximately 40 deposit locations were used. The local conditions of deposit bins varied, and climate-controlled locations were preferred, but the temperatures of deposited samples were not regulated. Participants could drop-off samples 24 hours a day (dependent on location accessibility), and samples were usually collected twice daily.

Sample registration occurred when a participant electronically registered their collection tube ID to a custom HIPAA-compliant database and sample management soft-ware. Sample registration required participants to enter a 10-digit numeric tube ID and 5-digit alphanumeric sequence. The alphanumeric sequence was derived from the 10-digit sequence, so successful registration required both sequences to be correctly entered, thus minimizing human input error. Saliva samples had nucleic acids extracted and assayed for SARS-CoV-2 infection by RT-PCR. From April 2020 to October 2021, all collected saliva sample extracts were assayed using the TaqPath COVID19 Combo Kit. After October 2021, saliva samples extracts were first analyzed using the TaqPath COVID-19 2.0 assay for diagnosis, and positive samples had their nucleic acids extracted and assayed using the TaqPath COVID19 Combo Kit assay. Sample results were electronically returned after appropriate approval by quality control and assurance officers under CLIA compliant protocols.

### 2.6 Participant demographics curation

During online sample registration, participants were asked to self-identify their sex, race, and ethnicity. Participants were also asked to enter their birthday. For each participant’s sex, race, and ethnicity, categorization of a participant was based on concordance of all of a patient’s registered samples. To be included as part of a cohort, all of a participant’s samples must have been registered with the same categorization (i.e., Male, Female, Unknown), or provided no response on other sample registrations. If a conflicting answer was provided on at least one tube, then the participant was assigned ‘Invalid or no answer’ for that categorization. For patient age at first test, the birthdate entered on the first registered sample was used to calculate the participant’s age. Ages exceeding 113 years were assigned ‘Invalid or no answer.’

### 2.7 Next generation sequencing of SARS-CoV-2

Sequencing facilities were in the same building where the diagnostic testing was performed. NGS libraries were built on nucleic acid extracts using the COVIDSeq Test (Illumina, San Diego, CA, USA) using ARTICv3, ARTICv4, or ARTICv4.1 [16] primer sets. Libraries were prepared for sequencing using a custom automation method on a Biomek i*7* liquid handler. Libraries were sequenced on an Illumina NextSeq2000 using 2 × 109 paired end reads. Reads were quality trimmed, filtered, and mapped using a custom pipeline using default parameters on software algorithms up to date at the time of analysis unless noted. Reads were trimmed of adapter sequences using trim-galore [20] (a wrapper for Cutadapt [21] and FastQC [22]) and aligned to the Wuhan1 reference genome (Genbank MN90894*7*.3) using the Burrows-Wheeler aligner, BWA-MEM [23]. Primer sequences were removed from reads using iVAR (arguments: -e, -q 30) [24] software. Consensus genomes were created using samtools (arguments: -t 0.60, -m 10, -n N) [25], given lineage assignments using pangolin [26], and validated using VADR (arguments: --glsearch, -s, -r, --nomisc, --lowsim5seq 2, --lowsim5ftr 2, --lowsim3seq 2, --lowsim3ftr 2, --alt_pass peptrans, --alt_fail lowscore,fstukcft,insertnn,deletinn) [27].

### 2.8 Statistical analysis

LOWESS/LOESS regression and Pearson correlation were calculated using GraphPad Prism software (Insight Partners, New York City, New York, USA). Ellipses generation and Wilson’s confidence interval calculation was performed using R software.

## 3. Results

### 3.1 Saliva is a stable and reliable sample matrix for SARS-CoV-2 testing

To test the feasibility of using saliva as a SARS-CoV-2 sample collection and extraction matrix, we first assayed the limit of detection on pooled, SARS-CoV-2 negative saliva samples with known quantities of spiked-in, heat inactivated SARS-CoV-2 virus particles (**Table 1**). Using a preliminary sample size of 3 samples per concentration, we obtained positive results for SARS-CoV-2 in all 3 samples down to 12.2 virus particles/µL. In order to confirm using 12.5 as our lower limit of detection, we expanded the sample size from 3 to 24 samples for a series of low virus concentrations. We successfully detected SARS-CoV-2 in all samples (24/24) at 1x, 2x, 4x, 8x, 16x, and 32x the lower limit of detection (**Table 1**). The ORF1ab and S gene loci appeared to be slightly more sensitive than the N gene locus, with consistently lower Ct values and detecting SARS-CoV-2 down to 6.1 particles/µL in one sample. Limit of detection analysis of NPSs resulted in similar detection patterns and a lower limit of detection of 1 virus particle per microliter of saliva (**Supplementary Table 1**).

**Table 1.** Limit of detection for SARS-Cov-2 virus particles in a saliva matrix.

| Assay | Virus particles/μL saliva | Positive/<br>Total | RNase P | Average Ct value (St. dev.) |  |  |
| --- | --- | --- | --- | --- | --- | --- |
|  |  |  |  | ORF1ab | N gene | S gene |
| Limit of detection (LOD) | 100000 | 3/3 | 21.81 (0.1) | 18.31 (0.16) | 18.84 (0.17) | 18.36 (0.12) |
|  | 50000 | 3/3 | 21.76 (0.1) | 19.46 (0.17) | 19.88 (0.06) | 19.44 (0.12) |
|  | 25000 | 3/3 | 21.7 (0.22) | 20.45 (0.22) | 20.75 (0.06) | 20.34 (0.13) |
|  | 12500 | 3/3 | 21.71 (0.19) | 21.4 (0.2) | 21.67 (0.18) | 21.29 (0.09) |
|  | 6250 | 3/3 | 21.72 (0.19) | 22.14 (0.35) | 22.55 (0.12) | 22.01 (0.35) |
|  | 3125 | 3/3 | 21.61 (0.13) | 23.11 (0.16) | 23.56 (0.19) | 23.13 (0.21) |
|  | 1562.5 | 3/3 | 21.65 (0.04) | 24.16 (0.2) | 24.46 (0.08) | 24.15 (0.18) |
|  | 781.3 | 3/3 | 21.63 (0.2) | 25.28 (0.36) | 25.42 (0.19) | 25.3 (0.27) |
|  | 390.6 | 3/3 | 21.68 (0.12) | 25.83 (0.2) | 26.39 (0.1) | 25.89 (0.18) |
|  | 195.3 | 3/3 | 21.67 (0.11) | 27.04 (0.11) | 27.48 (0.1) | 27.25 (0.07) |
|  | 97.7 | 3/3 | 21.53 (0.19) | 27.59 (0.27) | 28.38 (0.16) | 27.84 (0.48) |
|  | 48.8 | 3/3 | 21.53 (0.1) | 28.35 (0.57) | 29.93 (0.81) | 29.76 (0.92) |
|  | 24.4 | 2/3 | 21.49 (0.25) | 30.9 (2.39) | 31.99 (0.82) | 31.3 (0.77) |
|  | 12.2 | 3/3 | 21.5 (0.39) | 31.68 (0.48) | 37.3 (0.82) | 35.49 (2.09) |
|  | 6.1 | 1/3 | 21.62 (0.04) | 35.15 | - | 36.65 |
|  | 3.1 | 0/3 | 21.78 (0.21) | - | - | - |
| Confirmation | 12.5 (1x LOD) | 24/24 | 22.89 (0.22) | 30.36 (0.61) | 30.78 (0.49) | 30.54 (0.75) |
|  | 25 (2x LOD) | 24/24 | 22.71 (0.26) | 29.35 (0.54) | 29.83 (0.55) | 29.38 (0.73) |
|  | 50 (4x LOD) | 24/24 | 22.68 (0.36) | 28.03 (1.3) | 28.5 (1.36) | 28.08 (1.43) |
|  | 100 (8x LOD) | 24/24 | 22.48 (0.7) | 26.91 (1.49) | 27.65 (0.71) | 26.8 (2.17) |
|  | 200 (16x LOD) | 24/24 | 22.85 (0.19) | 26.25 (0.51) | 26.89 (0.24) | 26.08 (0.54) |
|  | 400 (32x LOD) | 24/24 | 22.78 (0.29) | 25.14 (0.92) | 26.01 (0.38) | 25.03 (0.88) |

With self-collection there are often delays between saliva collection and sample processing, so we tested SARS-CoV-2 virus particle detection at different times up to *7*2 hours at four storage temperatures (**Figure 1**). At the ORF1ab locus, Ct values deviated by less than 1 Ct over *7*2 hours for the −20°C, 4°C, and 20°C samples. There were greater deviations observed in samples incubated at 42°C, but differences to the initial Ct values stayed within 2 Ct values. Slightly greater Ct deviations were observed at the N gene and S gene loci. Overall, the differences we observed at the −20°C and 4°C temperatures deviated less than those at incubated at 20°C, and the samples incubated at 42°C showed the greatest deviations.

**Figure 1.**
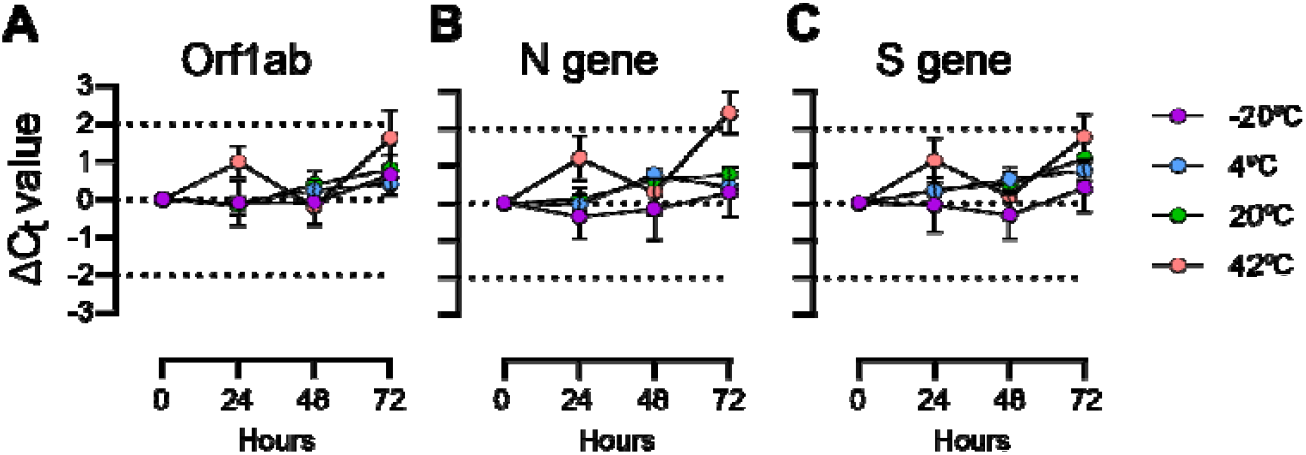
TaqPath Ct differences for saliva samples with spiked-in SARS-CoV-2 particles incubated at four times over *7*2 hours. (a) Orf1ab gene primer/probe target; (b) N gene primer/probe target; (c) S gene primer/probe target.

Finally, we wanted to compare the concordance of saliva-based SARS-CoV-2 testing to ‘gold-standard’ NP swab SARS-CoV-2 testing (**Table 2**). We tested 148 paired NP and saliva samples for SARS-CoV-2 using the TaqPath Combo Kit assay. There was 95% positive concordance (19/20 samples; 95% Wilson’s CI: *7*6.4%-99.1%) and 99.2% negative concordance (12*7*/128 samples; 95% Wilson’s CI: 95.*7*%-99.9%) between the two sampling methodologies.

**Table 2.**
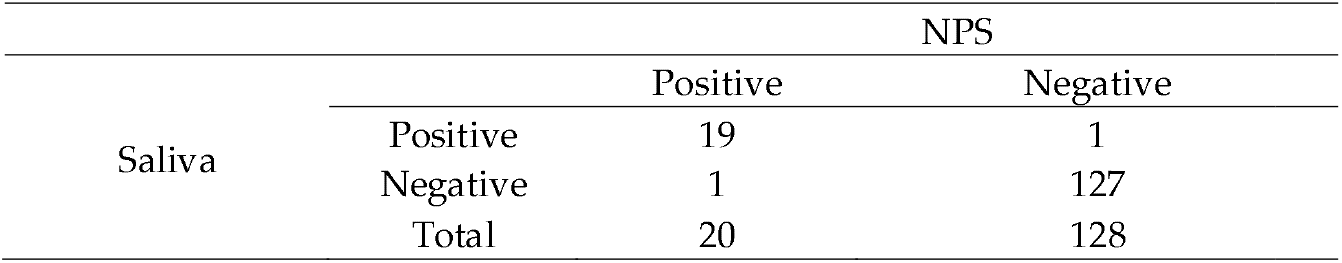
Comparison of nasopharyngeal swab (NPS) and saliva sampling method for detection of SARS-CoV-2 infection using the TaqPath COVID-19 Combo Kit qPCR assay.

With the finding that SARS-CoV-2 could be stably detected in saliva with good concordance to NP swab testing, we continued with large-scale self-administered saliva collection and RT-PCR SARS-CoV-2 surveillance.

### 3.2 Participant demographics of saliva-based COVID-19 surveillance

From April 2, 2020, to December 31, 2023, 14348*7*3 SARS-CoV-2 RT-PCR assays were performed on participant-collected saliva samples (**Table 3**). Over the surveillance period, 94,330 (6.6%) samples tested positive for SARS-CoV-2 and 10,922 (0.8%) samples were invalid. Most samples were collected from three Arizona counties: Maricopa County (*7*4.5%), followed by Coconino County (15.8%), and Pima County (6.3%), with the remaining samples (3.2%) coming from other counties. There were 366,681 unique participants who provided saliva samples. Slightly more participants identified as male (50.*7*%) than female (48.2%). Most participants identified as white (55.2%), but a large portion of participants provided an invalid answer or no answer (16.2%). A sizable portion of participants identified Hispanic or Latino ethnicity (21.9%).Most participants were aged 18-21 (21%) at the time of their first collection, indicating that college-aged students have a high representation in the participant set.

**Table 3.**
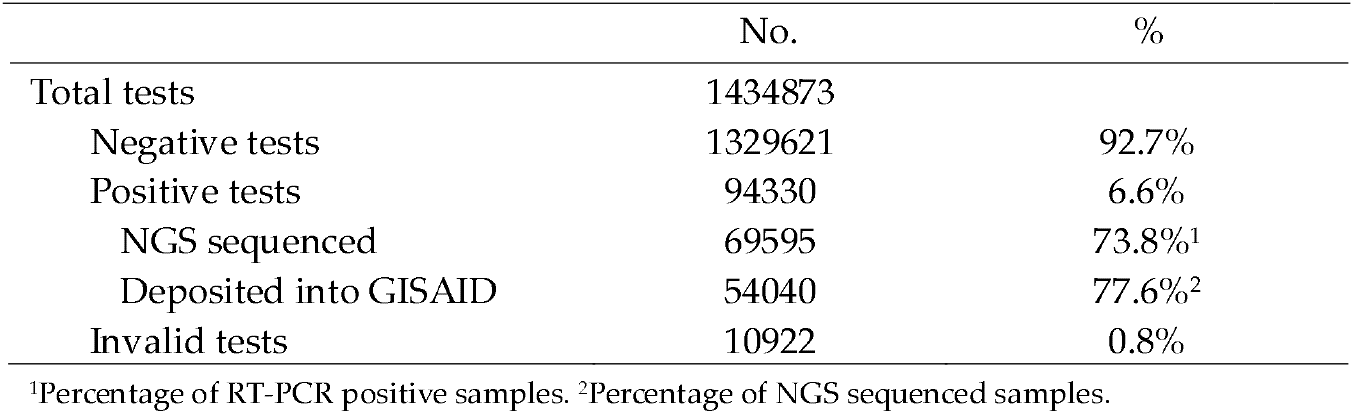
SARS-CoV-2 RT-PCR and NGS assays performed from April 2020 to December 2023.

|  |  |  |
| --- | --- | --- |
| Total tests | 1434873 |  |
| Negative tests | 1329621 | 92.7% |
| Positive tests | 94330 | 6.6% |
| NGS sequenced | 69595 | 73.8%<sup>1</sup> |
| Deposited into GISAID | 54040 | 77.6%<sup>2</sup> |
| Invalid tests | 10922 | 0.8% |

<sup>1</sup>Percentage of RT-PCR positive samples. <sup>2</sup>Percentage of NGS sequenced samples.

**Table 4.**
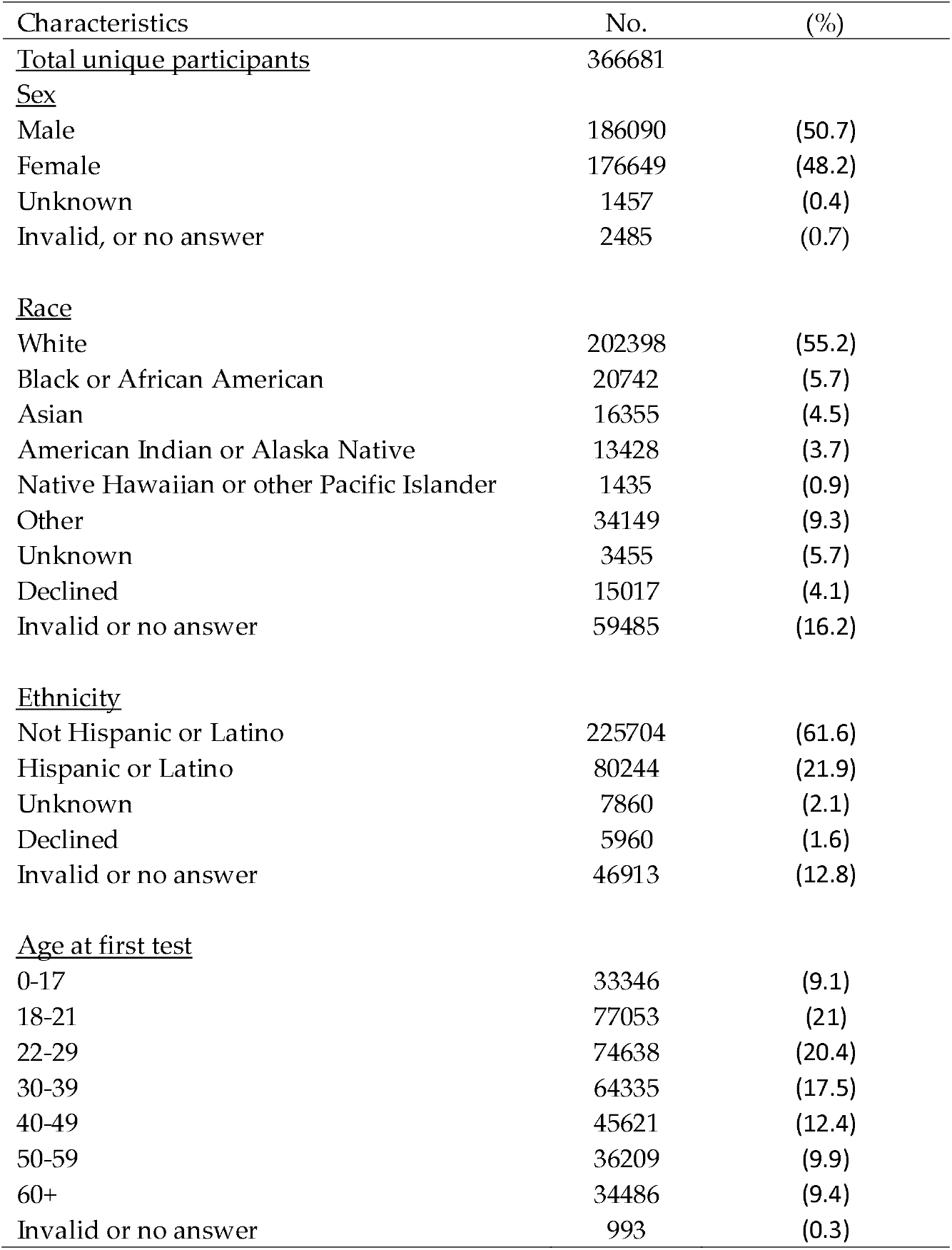
Demographics of surveillance participants from April 2020 to December 2023.

| Characteristics | No. | (%) |
| --- | --- | --- |
| <u>Total unique participants</u> | 366681 |  |
| <u>Sex</u> |  |  |
| Male | 186090 | (50.7) |
| Female | 176649 | (48.2) |
| Unknown | 1457 | (0.4) |
| Invalid, or no answer | 2485 | (0.7) |
| <u>Race</u> |  |  |
| White | 202398 | (55.2) |
| Black or African American | 20742 | (5.7) |
| Asian | 16355 | (4.5) |
| American Indian or Alaska Native | 13428 | (3.7) |
| Native Hawaiian or other Pacific Islander | 1435 | (0.9) |
| Other | 34149 | (9.3) |
| Unknown | 3455 | (5.7) |
| Declined | 15017 | (4.1) |
| Invalid or no answer | 59485 | (16.2) |
| <u>Ethnicity</u> |  |  |
| Not Hispanic or Latino | 225704 | (61.6) |
| Hispanic or Latino | 80244 | (21.9) |
| Unknown | 7860 | (2.1) |
| Declined | 5960 | (1.6) |
| Invalid or no answer | 46913 | (12.8) |
| <u>Age at first test</u> |  |  |
| 0-17 | 33346 | (9.1) |
| 18-21 | 77053 | (21) |
| 22-29 | 74638 | (20.4) |
| 30-39 | 64335 | (17.5) |
| 40-49 | 45621 | (12.4) |
| 50-59 | 36209 | (9.9) |
| 60+ | 34486 | (9.4) |
| Invalid or no answer | 993 | (0.3) |

### 3.3 Self-administered saliva collection surveillance reflects community COVID19 epidemiology

Testing frequency was high during the initial surveillance period through 2022 but was greatly diminished throughout 2023 (**Figure 2A**). Generally, total testing was low in the summer months and higher during the academic school year. Test administration peaked in December 2020 and experienced another large rise in December 2021. Po**s**itivity rate fluctuated over the surveillance period, with peak positivity during January **2**022. Interestingly, we observed that after July 2022, the positivity rate remained elevated compared to previous rates.

**Figure 2.**
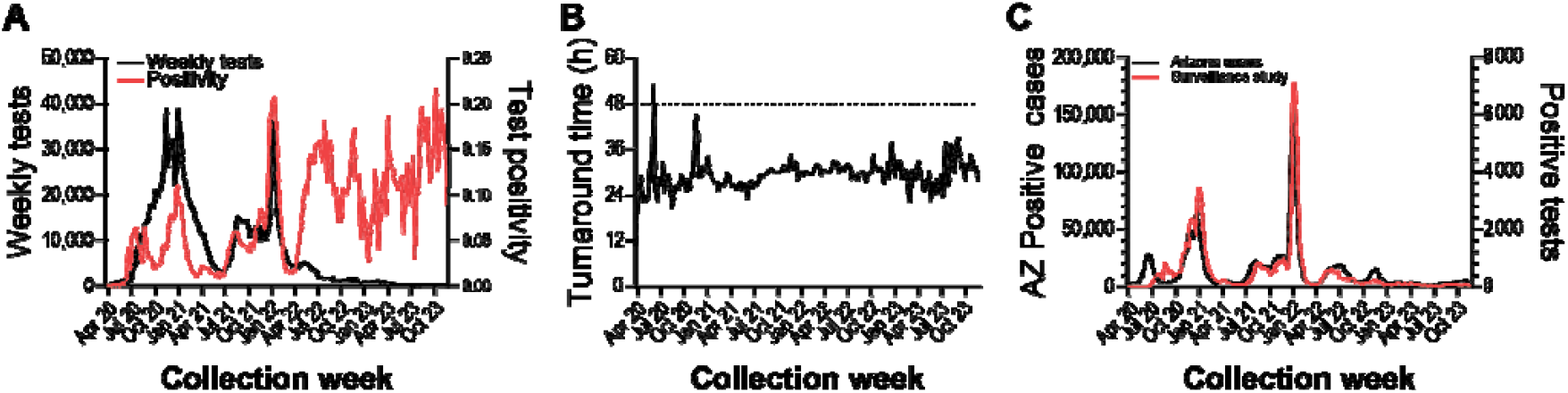
SARS-CoV-2 RT-PCR testing surveillance from April 2020 to November 2023. (**a**) Left axis: Weekly tests performed. Right axis: Weekly test positivity. (**b**) Average weekly test result turnaround time. (**c**) Left axis: Weekly Arizona SARS-CoV-2 case count. Right axis: Surveillance weekly positive samples.

The median turnaround time from participant sample registration to electronic delivery of RT-PCR results was 29.2 hours, and weekly average turnaround times rarely exceeded 48 hours (**Figure 2B**). We wanted to determine if our surveillance strategy and response time was indicative of SARS-CoV-2 infection in the broader community, so we compared our weekly positive test counts to Arizona COVID19 case counts (**Figure 2C**). We found that trends between the two metrics were highly correlated (Pearson r = 0.90*7*3, 95% CI: 0.88-0.93, p < 0.0001). Taken together, we observed that the diversity and responsiveness of our surveillance testing strategy sufficiently captured the local region’s epidemiological status.

### 3.4 Randomly selected test participation detected infection outbreaks

Our surveillance program was comprised of community participants self-initiating testing as well as participants invited to test by an affiliate organization (e.g. school, employer), so we wanted to compare how positivity between self-initiated testing compared to a population randomly selected and invited to test. Through May 2020 and February 2022, we compared test positivity for an organization implementing randomized, invited testing to individuals self-initiating testing from the same geographic area. We observed that test positivity for self-initiated testing was generally higher than invited testing, but followed similar upward and downward trends (**Figure 3A**).

**Figure 3.**
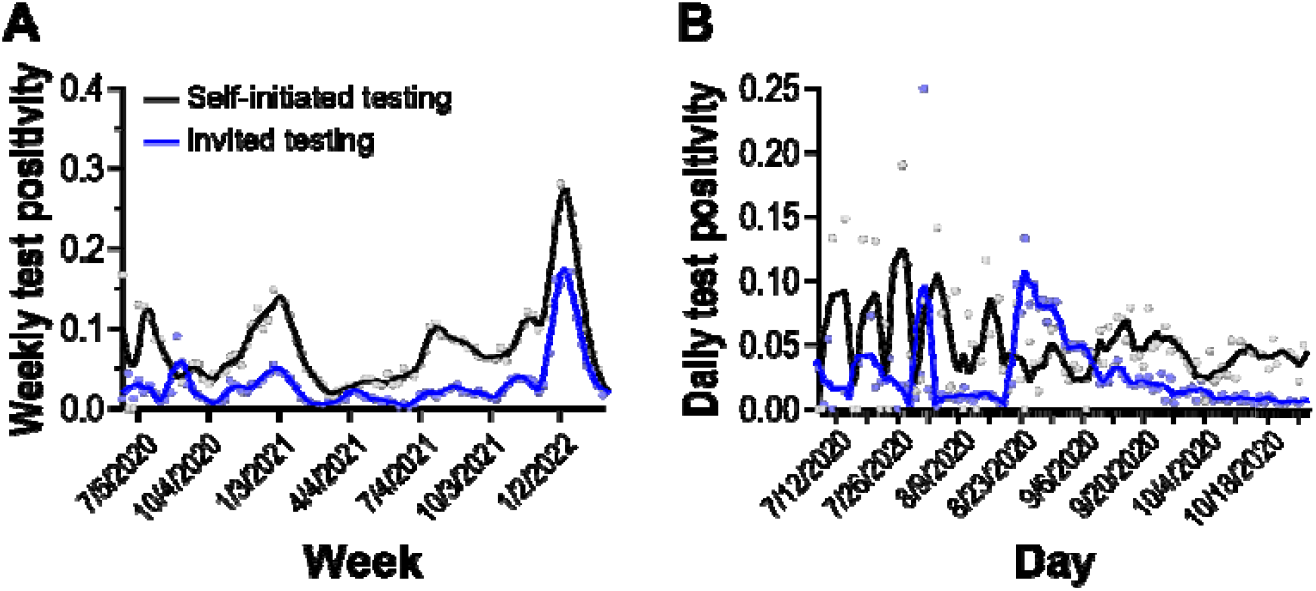
Test positivity rate of self-initiated and invited testing cohorts. Solid lines are 2^nd^ order smoothing functions across 3 neighbor data points. (**a**) Weekly test positivity during the surveillance period. (**b**) Daily test positivity around outbreak period (see text).

Starting on August 22, 2020, we observed a period of 1*7* days where the invited testing cohort displayed test positivity higher than the self-initiated testing cohort (**Figure 3B**). It took approximately 40 days for test positivity in the randomly selected invited cohort to return to pre-elevated levels. Overall, we observed that randomly selected invited testing can detect outbreaks not captured in self-initiated sampling strategies.

### 3.5 Saliva sample collection enabled whole viral genome sequencing of clinical positives for genomic epidemiology investigations

In order to monitor for the introduction of VOCs and the emergence of novel mutations within circulating variants, whole viral genome sequencing of RT-PCR positive participant samples was performed. Over the surveillance period, sequencing of 69,595 saliva samples was attempted and 54,040 consensus genome sequences received sufficient coverage for PANGO designation and were submitted to the GISAID database (**Table 3**).

From September 2020 through February 2021, saliva and nucleic acid extracts from SARS-CoV-2 positive samples were stored at −80°C until sequencing infrastructure was finalized. When NGS started in February 2021 through December 2021, weekly GISAID submissions generally matched RT-PCR positive sample collections (**Figure 4A**). In late December 2021 to February 2022, a surge of positive RT-PCR samples caused sample influx to exceed sequencing capacity. Over this period, samples collected on the most recent day were prioritized for WGS and others were banked for later sequencing. From February 2022 to May 2022, GISAID submissions greatly surpassed sample collections. During this period, banked samples were supplemented into NGS runs to meet sequencing capacity. After May 2022, GISAID submissions tracked sample collections and overall sample collections decreased compared to earlier dates.

**Figure 4.**
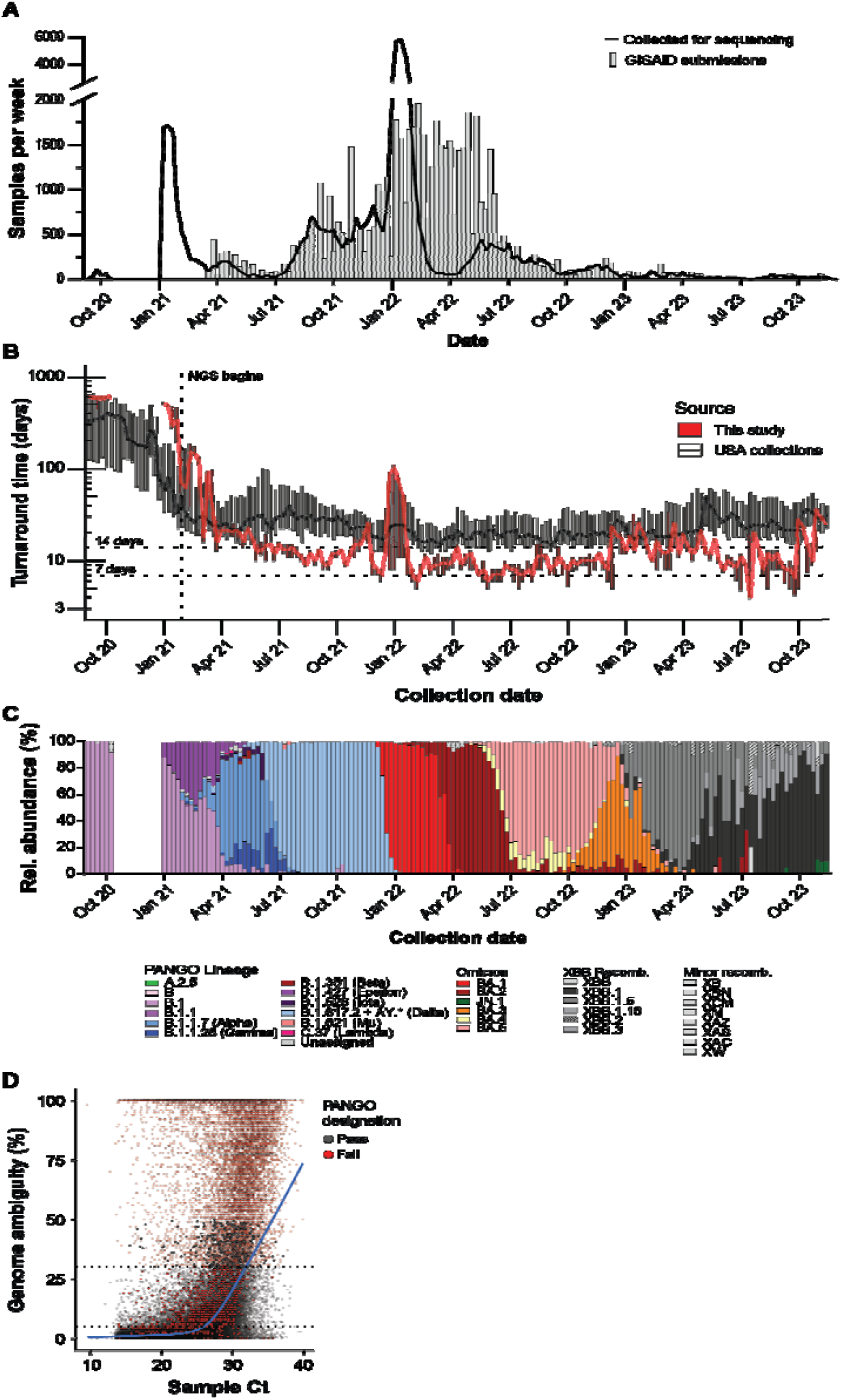
Whole viral genome sequencing of SARS-CoV-2 positive saliva samples. (**a**) Weekly RT-PCR positive samples (line) and weekly GISAID submissions (bars). (**b**) GISAID collection to submission turnaround time for surveillance program and other USA samples. Solid line corresponds to median turnaround time. Bottom and top of boxes correspond to Q1 and Q3 values, respectively. (**c**) Relative weekly abundance of SARS-CoV-2 PANGO lineages. (**d**) TaqPath Ct values and consensus genome ambiguity of saliva samples. Blue line shows LOWESS smoothing of all samples. (**b**)

Sample sequencing turnaround time, the time between participant sample registration and GISAID submission, is important to maintaining a ‘real-time’ analysis of circulating variants, therefore we analyzed the sample turnaround for our sequencing samples [28]. Turnaround time fluctuated over the surveillance period (**Figure 4B**). Samples collected and banked before sequencing infrastructure was established in February 2021, had the longest turnaround times. In May 2021, median turnaround times shortened to a desired *7*-14 day turnaround time. Median turnaround times increased during December values remained within the *7*-14 day window. When we compared our sequencing turnaround times to other sequencing laboratories in the USA, we observed that our median turnaround times were usually lower than weekly national medians. Fo our study and for other USA samples, 50% of weekly median turnaround times were below 12 and 23 days, respectively **(Figure 4B)**.

In order to observe trends in SARS-CoV-2 virus evolution, we looked at the relative abundances of each lineage using the GISAID lineage calls from our submitted samples over the surveillance period **(Figure 4C)**. Major lineages reaching a majority relative abundance (>=50% of samples) in any week over the surveillance period were: B.1, B.1.1.*7* (Alpha), B.1.61*7*.2 (Delta, with AY sublineages), BA.1 (Omicron), BA.2 (Omicron), BA.3 (Omicron), BA.5 (Omicron), XBB.1 (Omicron-recombinant), XBB.1.5 (Omicron-recombinant), XBB.1.16 (Omicron-recombinant). Notable lineages that were detected but did not meet majority abundance were: A.2.5, B.1.1.28/P.1 (Gamma), B.1.351 (Beta), B.1.526 (Iota), B.1.621 (Mu), BA.4 (Omicron), C.3*7* (Lambda), and JN.1 (Omicron). The following non-XBB.1 recombinant lineages were detected during the surveillance period: XAC, XAS, XAZ, XB, XBB.2, XBB.3, XBN, XCM, XM, XW.

Finally, to assess the effect of viral load on sequencing performance, we compared TaqPath C_t_ values to consensus genome ambiguity **(Figure 4D)**. We observed that fewer ambiguities were obtained at higher viral loads and ambiguity frequency rose as viral load decreased. Performing a LOWESS regression of all samples found that C_t_ values below 32.0 and 26.0 were associated with fewer than 30% and 5% ambiguities, respectively. We observed a low frequency (n = 998, 1.*7*%) of genomes with less than 30% ambiguity that failed pangolin lineage designation or GISAID acceptance. We also observed that a low proportion (n= 342, 0.6%) of consensus genomes with over 30% ambiguities were able to receive pangolin lineage designations and GISAID acceptance.

### 3.6 Diagnostic test failure allowed rapid genotyping of saliva samples

The TaqPath Combo Kit detects the presence of SARS-CoV-2 genomes using primers and probes with homology to the ORF1ab, N, and S genes. When we compared Orf1ab gene or N gene Ct values to S gene Ct values, we observed populations of genomes with discordant S gene Ct values **(Figure 5A)**. Genome sequencing revealed that this population was primarily composed of specific lineages. Of the major, notable, and recombinant lineages observed over the surveillance period **(Figure 4C)**, we observed significantly large differences in median TaqPath Ct values for B.1.1.*7* (Alpha), BA.1 (Omicron), BA.3 (Omicron), BA.4 (Omicron), BA.5 (Omicron), JN.1 (Omicron), XAS, XAZ, and XM lineages compared to the B.1.1 lineage (**Figure 5B**; Tukey HSD, p < 0.005). Median Orf1ab Ct values differed between lineages but were not large enough to account for the observed differences **(Supplementary Figure S1)**. The lineages displaying S gene target failure (SGTF) all contain the H69del/V*7*0del mutations.

**Figure 5.**
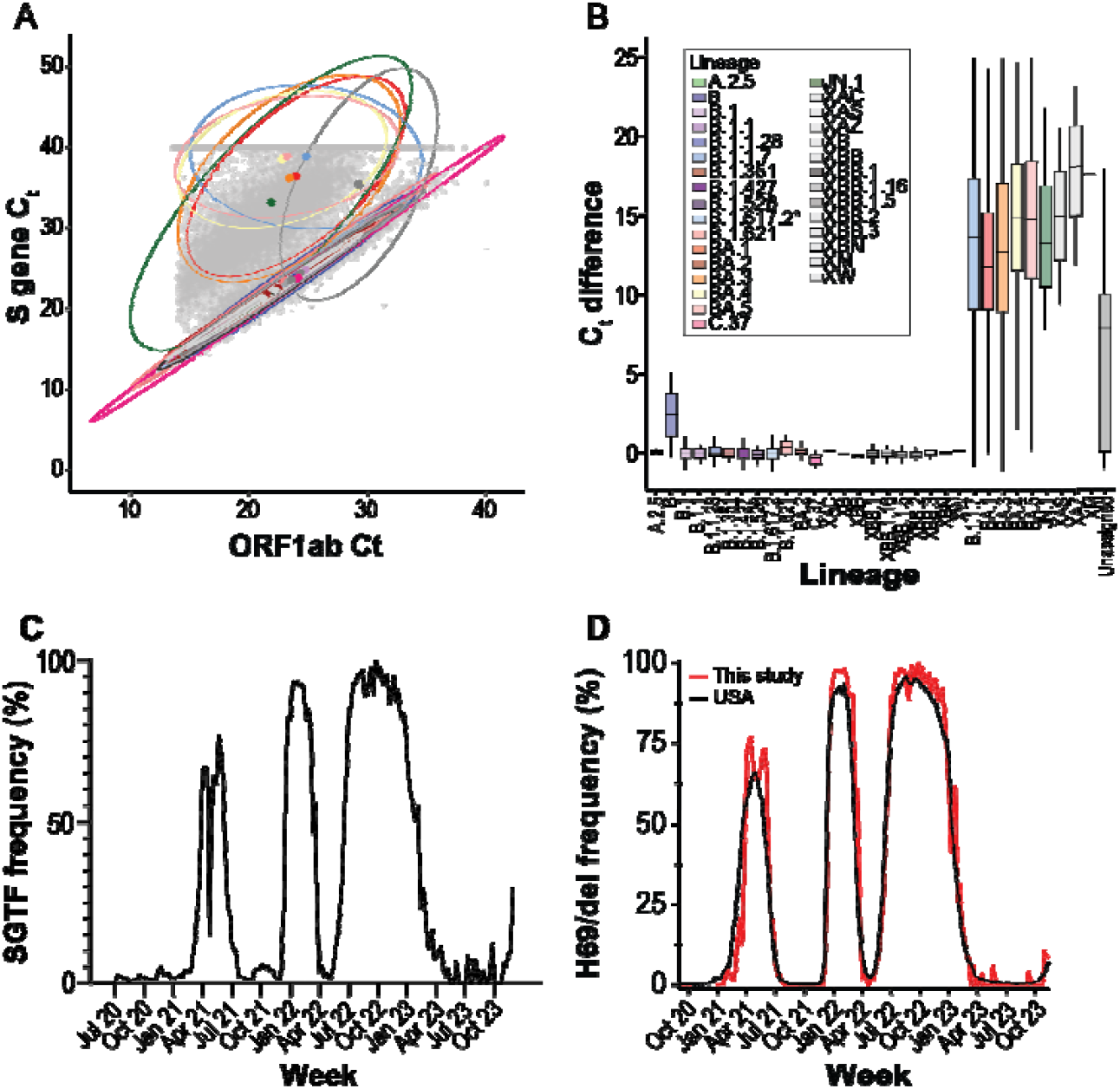
S gene target failure (SGTF) on TaqPath COVID19 Combo Kit RT-PCR assays. (**a**) Mean ORF1ab/N gene Ct values and S gene Ct values of positive saliva samples. Ellipses capture 95% of samples belonging to each PANGO variant. Colored points indicate ellipse centroid. Lineage coloration is shared between panels (a) and (b). (b) Difference between S gene and mean ORF1ab/N gene Ct values for PANGO variants displaying SGTF. The bottom, middle, and top of boxes correspond to Q1, Q2, and Q3, respectively. Box whiskers correspond to the largest value no further than 1.5*IQR. (**c**) Weekly SGTF prevalence in local samples. (**d**) Weekly abundance of samples containing S:H69del mutation in USA and local samples.

We detected three periods of high SGTF frequency over the surveillance period, as well as growing frequency at the end of the survey period (Figure 5C). We compared the frequency of our sequenced genomes containing H69/V*7*0 deletion to the frequency of other USA genomes and observed that the mutation frequency patterns were identical to SGTF frequency (Figure 5D). In summary, multiple times during the surveillance period, RT-PCR diagnostic test failure enabled rapid H69/V*7*0 deletion genotyping and facilitated a fast way to monitor changing lineage abundance.

## 4 Discussion

In this study we report the multi-year effort of monitoring the introduction and evolution of SARS-CoV-2 in Tempe, Arizona. Self-collected saliva samples were an adequate matrix for qualitative diagnosis of SARS-CoV-2 infection, and we obtained robust genome sequencing from most positive samples. Using our surveillance system, we as-sessed the epidemiological health of the community and monitored evolution of the SARS-CoV-2 virus throughout the pandemic.

Our multi-year surveillance was primarily located in an urban metropolitan area of a major US city. Due to the high testing volume, high sequencing depth, and fast turnaround times, our surveillance program provided a robust representation of SARS-CoV-2 epidemiology [28]. Variant introduction and abundance patterns that we observed concur with patterns seen in other regional studies [29]. Upward and downward trends in testing frequency and positivity also appear consistent with regional (Figure 1C) and national [30] case counts and test positivity. However, in this study, we observe an overall increase in test positivity dynamics before and after infections peaked during the Omicron BA.1 variant introduction in January 2022 **(Figure 2A)**.

We also observed periods where self-initiated testing was higher than invited testing **(Figure 3)**. Patterns of test positivity might be attributed to the motivations behind SARS-CoV-2 self-initiated testing. Testing motivations are multifaceted, involving personality, behavioral, clinical, and economic factors [31,32]. The availability of SARS-CoV-2 vaccines after December 2020 [33] may be also have contributed to altered testing motivations. Seroprevalence surveillance during this time period, showed a large number of adults had antibodies to SARS-CoV-2 through vaccination, infection, or both [34]. Attitudes to testing may have been altered due to perceived protection status from vaccination or naturally acquired immunity (i.e., recent infection). Finally, the increased availability of commercial, at-home, SARS-CoV-2 tests may have altered the surveillance population and their motivations. Our study shows that while self-initiated and invited testing are generally in concordance, further understanding of testing motivations and how to promote self-initiated testing may improve the representativeness of self-initiated testing.

Co-localizing RT-PCR and WGS operations resulted in valuable benefits. Close physical proximity reduced sample transit and storage time, minimized the number of freeze-thaw cycles a sample was exposed to, and facilitated communication between teams. Shipping time, storage time, and freeze-thaw cycles are all factors that negatively affect RNA stability [35]. Enhanced communication and coordination abilities allowed catching and correcting errors, as well as assisted in coordinating sequencing priority based on SGTF results.

In addition to internal communication, external guidance by state and federal agencies provided valuable information. The US Food & Drug Administration guidance documents assisted in the development of the Arizona Biodesign Clinical Testing Lab and establishing our surveillance program [36]. Federal agencies also organized regular status and discussion panels, such as the CDC’s Sequencing for Public Health Emergency Response, Epidemiology, and Surveillance (SPHERES) consortium [37]. These supplementary communication networks facilitated prompt dispersal of information between testing sites, such as when novel variants displayed reduced sensitivity to diagnostic assays.

The genetic determinants of SGTF were known soon after introduction of the Alpha variant [12]. The H69del/V*7*0del mutation in the Spike protein provides no immune escape properties itself but rescues other Spike protein mutations with increased antibody evasion but impaired infectivity [38]. The increased prevalence of SGTF on SARS-CoV-2 specimens led to the design and adoption of TaqPath 2.0 for primary diagnostic use. However, the continued use of the TaqPath Combo Kit Assay continued to provide important public health data. Broadly, it allowed facilities without sequencing capabilities to monitor variant introduction [39]. SGTF provided fast variant typing during the Alpha-Delta, Delta-Omicron BA.1, Omicron BA.1-Omicron BA.2, Omicron BA.2-Omicron BA.3/4/5, and Omicron BA.3/4/5-XBB transitions **(Figures 4C and 5A)**. For sequencing laboratories, SGTF was useful when sample positivity exceeded sequencing capacity, as it allowed researchers to direct sequencing efforts towards samples suspected of being newly introduced variants. Subsequent WGS allowed confirmation of those variants and allowed surveillance for novel mutations within introduced lineages.

Although most samples with low genome ambiguity were accepted for database submission, we observed a small number (n = 9*7*9, 1.4%) of samples that had high genome coverage (>30%) but failed lineage designation or GISAID submission **(Figure 4D)**. Within our workflow, submission of consensus genomes to the GISAID database was conditional upon assignment of a PANGO lineage by pangolin software at the time of sequencing analysis. Further, samples with contamination or true co-infection may have received mixed-lineage designations and been excluded from database submission. There may also be some cases where a sample contained ambiguities at nucleotides critical in lineage designation or samples belonged to undesignated lineages at the time of analysis. Amplicon-based sequencing was the primary sequencing method, therefore mutations in viral genomes located within primer binding regions would cause reduced sequencing coverage of that amplicon. This phenomenon, termed amplicon drop-out, has occurred multiple times over the course of SARS-CoV-2 evolution [40,41]. Samples with ambiguities in positions critical to lineage defining specificity may have been incorrectly assigned a lineage or left unassigned, causing them to fail submission. During our study, we obtained genome completeness scores, with respect to RT-PCR Ct values, comparable to those observed in another study [42], illustrating that saliva is a viable medium for robust, large scale genomic surveillance.

Ultimately, in this study we determined that self-collected saliva-based surveillance assays can provide important epidemiological information during a public health crisis. We observed that our surveillance system (i) provided an effective means to broadly assess community health and epidemic status, (ii) provided a means to effectively detect outbreaks within the population to target remediation strategies, and (iii) quickly provided high-quality material to enable the monitoring of pathogen evolution by WGS during an epidemic crisis. Due to its simple collection and low invasiveness, continued examination of saliva as a testing matrix for routine and pandemic pathogen surveillance is important.

## Supplementary Materials

The following supporting information can be downloaded at: https://www.mdpi.com/article/doi/s1, Figure S1: TaqPath COVID-19 Combo Kit Orf1ab Ct values of saliva samples by SARS-CoV-2 lineages; Table S1: Limit of detection analysis of SARS-CoV-2 detection in nasopharyngeal swabs using the TaqPath COVID-19 Combo Kit qPCR assay.

## Author Contributions

Conceptualization, V.M., J.L., and E.S.L; methodology, T.R., V.M., I.S and E.S.L; software, I.S., and S.H; formal analysis, S.H and V.M; data curation, S.H.; writing—original draft preparation, S.H.; writing—review and editing, S.H., V.M., and J.L.; visualization, S.H.; supervision, C.C. and V.M.; project administration, V.M.; funding acquisition, V.M., E.S.L., and J.L. All authors have read and agreed to the published version of the manuscript.” Please turn to the CRediT taxonomy for the term explanation. Authorship must be limited to those who have contributed substantially to the work reported.

## Institutional Review Board Statement

Ethical review and approval were waived for this study due to the use of deidentified samples; metadata did not include any participant-identifiable information.

## Informed Consent Statement

Not applicable.

## Data Availability Statement

Viral genome sequences generated in this surveillance program were submitted to the GISAID nCov database and can be obtained using the EPI_SET ID: EPI_SET_250224ka. RT-PCR results, sequencing results, and patient metadata is available at DataDryad: https://doi.org/10.5061/dryad.z08kprrsh

## ASU Biodesign Clinical Testing Lab (ABCTL) Diagnostic and Sequencing Teams

(alphabetical listing): Ajeet Bains, Bradley Bobbett, Veronica Boyle, Sean Dudley, Alexis Emerson, Michael Fiacco, Izamer Garcia, Kristina Gonzalez-Wartz, Akshaya Gunasekaran, Valerie Harris, LaRinda A. Holland, Ching-Wen Hou, James C. Hu, Preston Hunter, Nathaniel Johnson, Emily A. Kaelin, Mark Knappenberger, Victoria R. Leonard, Kyle Lewis, Jessica Lukosus, Madhuranga Thilakasiri Madugoda Ralalage Don, Mitch Magee, Shodhan Manda, Rabia Maqsood, Winston Matthews, Aaron McDonald, Tianchen Mu, Advait Murugan, Nicholas Nabours, Benjamin Nussel, Yasmine Parra, Peter Patterson, Nghia C. Pham, Eric Reamer, Michael Ritchie, Tyler Schroeder, Amit Arunkumar Sharma, Peter T. Skidmore, Matthew F. Smith, Regan A. Sullins, Alexis Thomas, An Tieu, Kevin Tinnin, and Chau Tran, Lily I. Wu.

## Abbreviations

The following abbreviations are used in this manuscript:

(S)GTF (S): Gene Target Failure
CDC: Centers for Disease Control and Prevention
COVID-19: Coronavirus disease 2019
EUA: Emergency Use Authorization
GISAID: Global Initiative on Sharing All Influenza Data
HIPAA: Health Insurance Portability and Accountability Act
LOWESS/LOESS: Locally Weighted/Estimated Scatterplot Smoothing
NGS: Next-generation sequencing
NPA: nasopharyngeal aspirate
NPS: nasopharyngeal swab
PANGO(LIN): Phylogenetic Assignment of Named Global Outbreak (Lineages)
RT-PCR: reverse transcriptase polymerase chain reaction
SARS-CoV-2: Severe acute respiratory syndrome coronavirus 2
VOC: Variant of Concern
VOI: Variant of Interest
VUM: Variant Under Monitoring
WGS: Whole Genomes Sequencing
WHO: World Health Organization

