## Supplementary tables and figures for "Clinical Surveillance Identifies SARS-CoV-2 Outbreaks and Emergence of Novel Variants in Real-Time"

Table S1: Limit of detection analysis of SARS-CoV-2 detection in nasopharyngeal swabs using the TaqPath COVID-19 Combo Kit qPCR assay.

| Assay | Virus particles/µL saliva | Positive/Total | Average C_t_ value (St. dev.) | | | |
| --- | --- | --- | --- | --- | --- | --- |
|  |  |  | RNAse P | ORF1ab | N gene | S gene |
| Limit of detection (LOD) | 500 | 3/3 | 28.00 (0.22) | 21.73 (0.3) | 22.69 (0.28) | 19.34 (0.77) |
|  | 50 | 3/3 | 27.73 (0.57) | 25.55 (0.25) | 26.26 (0.19) | 22.77 (0.81) |
|  | 5 | 3/3 | 27.8 (0.07) | 26.85 (2.23) | 28.66 (0.88) | 25.27 (2.02) |
|  | 1 | 3/3 | 27.53 (0.24) | 33.16 (1.46) | 32.24 (0.7) | 32.87 (1.52) |
|  | 0.5 | 1/3 | 27.47 (0.03) | 33.26 | 34.12 | 33.72 |
|  | 0.25 | 0/3 | 26.87 (0.73) | - | - | - |
| Confirmation | 1 (1x LOD) | 24/24 | 24.51 (0.97) | 31.25 (3.31) | 31.37 (6.15) | 29.6 (5.04) |
|  | 2 (2x LOD) | 24/24 | 24.63 (0.15) | 31.22 (2.04) | 32.68 (1.08) | 28.74 (3.16) |
|  | 3 (3x LOD) | 24/24 | 24.13 (0.86) | 25.63 (2.89) | 29.49 (2.98) | 24.74 (3.65) |
|  | 5 (5x LOD) | 24/24 | 24.93 (0.14) | 29.11 (0.81) | 30.98 (0.62) | 26.13 (2.8) |

Supplementary Figure 1: TaqPath COVID-19 Combo Kit Orf1ab Ct values of saliva samples by SARS-CoV-2 lineages.

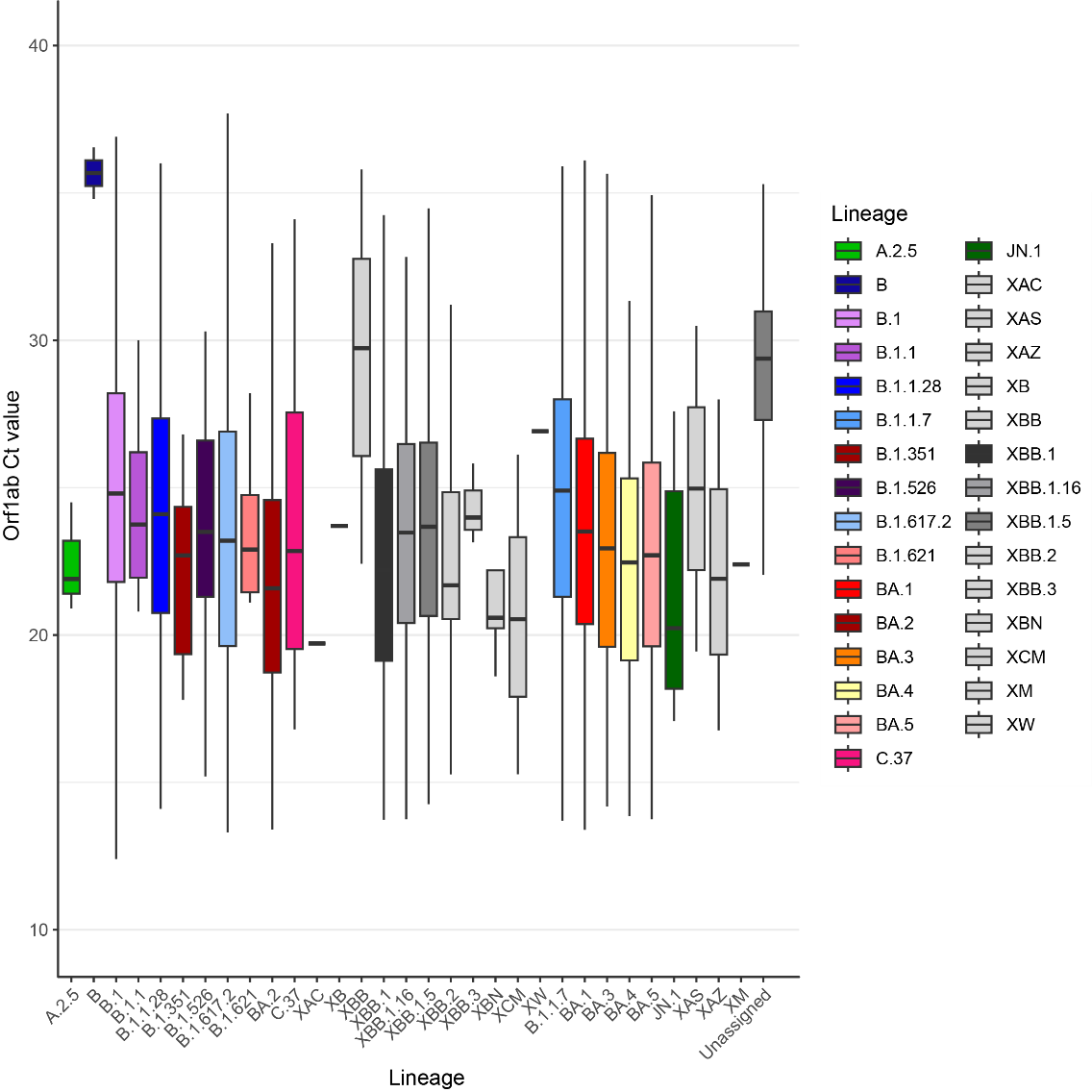
